# “Clinical characteristics, symptoms, management and health outcomes in 8,598 pregnant women diagnosed with COVID-19 compared to 27,510 with seasonal influenza in France, Spain and the US: a network cohort analysis”

**DOI:** 10.1101/2020.10.13.20211821

**Authors:** Lana Yin Hui Lai, Asieh Golozar, Anthony Sena, Andrea V. Margulis, Nuria Haro, Paula Casajust, Neus Valveny, Albert Prats-Uribe, Evan P. Minty, Waheed-Ul-Rahman Ahmed, Thamir M Alshammari, Daniel R. Morales, Heba Alghoul, Osaid Alser, Dalia Dawoud, Lin Zhang, Jose D. Posada, Nigam H. Shah, Clair Blacketer, Carlos Areia, Vignesh Subbian, Fredrik Nyberg, Jennifer C E Lane, Marc A Suchard, Mengchun Gong, Martina Recalde, Jitendra Jonnagaddala, Karishma Shah, Elena Roel, David Vizcaya, Stephen Fortin, Ru-fong Joanne Cheng, Christian Reich, George Hripcsak, Peter Rijnbeek, Patrick Ryan, Kristin Kostka, Talita Duarte-Salles, Daniel Prieto-Alhambra

## Abstract

**OBJECTIVES:** To describe comorbidities, symptoms at presentation, medication use, and 30-day outcomes after a diagnosis of COVID-19 in pregnant women, in comparison to pregnant women with influenza.

**DESIGN:** Multinational network cohort

**SETTING:** A total of 6 databases consisting of electronic medical records and claims data from France, Spain, and the United States.

**PARTICIPANTS:** Pregnant women with ≥ 1 year in contributing databases, diagnosed and/or tested positive, or hospitalized with COVID-19. The influenza cohort was derived from the 2017-2018 influenza season.

**OUTCOMES:** Baseline patient characteristics, comorbidities and presenting symptoms; 30-day inpatient drug utilization, maternal complications and pregnancy-related outcomes following diagnosis/hospitalization.

**RESULTS:** 8,598 women diagnosed (2,031 hospitalized) with COVID-19 were included. Hospitalized women had, compared to those diagnosed, a higher prevalence sof pre-existing comorbidities including renal impairment (2.2% diagnosed vs 5.1% hospitalized) and anemia (15.5% diagnosed vs 21.3% hospitalized).

The ten most common inpatient treatments were systemic corticosteroids (29.6%), enoxaparin (24.0%), immunoglobulins (21.4%), famotidine (20.9%), azithromycin (18.1%), heparin (15.8%), ceftriaxone (7.9%), aspirin (7.0%), hydroxychloroquine (5.4%) and amoxicillin (3.5%).

Compared to 27,510 women with influenza, dyspnea and anosmia were more prevalent in those with COVID-19. Women with COVID-19 had higher frequency of cesarean-section (4.4% vs 3.1%), preterm delivery (0.9% vs 0.5%), and poorer maternal outcomes: pneumonia (12.0% vs 2.7%), ARDS (4.0% vs 0.3%) and sepsis (2.1% vs 0.7%). COVID-19 fatality was negligible (N<5 in each database respectively).

**CONCLUSIONS:** Comorbidities that were more prevalent with COVID-19 hospitalization (compared to COVID-19 diagnosed) in pregnancy included renal impairment and anemia. Multiple medications were used to treat pregnant women hospitalized with COVID-19, some with little evidence of benefit. Anosmia and dyspnea were indicative symptoms of COVID-19 in pregnancy compared to influenza, and may aid differential diagnosis. Despite low fatality, pregnancy and maternal outcomes were worse in COVID-19 than influenza.

**WHAT IS ALREADY KNOWN ON THIS TOPIC:**

- Compared to non-pregnant women of reproductive age, pregnant women are less likely to experience typical COVID-19 symptoms, such as fever and myalgia.
- Obesity, high maternal age, and comorbid hypertension and diabetes are risk factors for severe COVID-19 among pregnant women.
- Despite relatively high rates of pneumonia and need for oxygen supplementation, fatality rates in pregnant women with COVID-19 are generally low (<1%).

**WHAT THIS STUDY ADDS:**

- Although not often recorded, dyspnea and anosmia were more often seen in pregnant women with COVID-19 than in women with seasonal influenza, in 6 databases from 3 countries (US, France, Spain).
- Renal impairment and anemia were more common among hospitalized than diagnosed women with COVID-19 during pregnancy.
- Despite limited data on benefit-risk in pregnancy, a large number of medications were used for inpatient management of COVID-19 in pregnant women: approximately 1 in 3 received corticosteroids (some may have been given for a pregnancy-related indication rather than for COVID-19 treatment), 1 in 4 enoxaparin, and 1 in 5 immunoglobulin, famotidine and azithromycin.
- Compared to influenza, there was a higher frequency of pregnancy-related complications (cesarean section and preterm deliveries), as well as poorer maternal outcomes (pneumonia, acute respiratory distress syndrome, sepsis, acute kidney injury, and cardiovascular and thromboembolic events) seen in pregnant women diagnosed with COVID-19.

## INTRODUCTION

As of 12^th^ October 2020, the coronavirus disease 2019 (COVID-19) pandemic has resulted in more than 37 million confirmed cases and more than 1 million deaths worldwide^1^. Latest data from the United States (US) Center for Disease Control and Prevention (CDC) showed that as of 24^th^ September 2020, a total of 23,222 cases of pregnant women with COVID-19 have been reported in the US alone^2^.

While the effects of COVID-19 on pregnancy and pregnant women are still not fully understood, studies based on other respiratory infections including influenza, pneumonia and middle-east respiratory syndrome (MERS) have indicated that changes in normal physiological and immunological functions, as well as hormonal levels during pregnancy may heighten the risk of severe illness^3-5^. Recent studies have shown that despite having relatively milder symptoms than the general population^6^, pregnant women with COVID-19 are at increased risk of hospitalization and admission to intensive care units^7^.

There are however, some unanswered issues of concern. First, there is a lack of evidence on whether pregnant women who were hospitalized had underlying conditions that made them more susceptible, as compared to those who were diagnosed with COVID-19 but not hospitalized. Second, the use of medications among pregnant women in the management of COVID-19 is also a topic of concern. While the efficacy and safety of many therapies are being actively evaluated in clinical trials, pregnant women are often excluded from these trials due to safety concerns, meaning there is a paucity of evidence around drug use in this vulnerable population. Third, with the simultaneous circulation of COVID-19 and seasonal influenza, it is also crucial to understand how the clinical manifestations and outcomes differ between COVID-19 and influenza patients to facilitate differential diagnosis and improve clinical management.

While the number of publications on COVID-19 in pregnancy has increased rapidly over time, most of these published studies are single center case reports, case series and observational studies with small sample sizes. A recent living systematic review among pregnant women with COVID-19 showed that of the 77 studies included, only 4 studies had a sample size of >1000^6^.

In our study, we aim to bridge existing gaps in the literature by describing the baseline socio-demographics, clinical characteristics, presenting symptoms, 30-day use of medications and outcomes of pregnant women diagnosed with and hospitalized for COVID-19. We also compare these with symptoms and outcomes seen amongst pregnant women diagnosed with influenza as a benchmark, in view of how many professional bodies are currently advocating pregnant women to receive their influenza vaccine with urgency^8, 9^.

## METHODS

### Study Design

The Characterizing Health Associated Risks, and Your Baseline Disease In SARS-COV-2 (CHARYBDIS) study is a large-scale multinational cohort study conducted across a network of hospital electronic health records (EHRs), primary care EHR, and health claims data standardized to the Observational Medical Outcomes Partnership (OMOP) Common Data Model (CDM). Through the Observational Health Data Sciences and Informatics (OHDSI) research network, the OMOP CDM supports large scale analytics and allows for generating robust and reproducible real-world evidence. Each participating institution retains their own data but makes it available for querying and statistical evaluation by locally running standardized analysis programs in a federated manner^10^. The study protocol for CHARYBDIS is available online^11^.

### Data Sources

From the 18 databases contributing data to the CHARYBDIS Study to characterize the history of COVID-19, only those with data on pregnant women with a clinical diagnosis of COVID-19 or a SARS-CoV-2 positive test were included in this analysis.

To be included in the study, databases had to have a minimum of 140 subjects in at least one of the COVID-19 cohorts. This cut-off was deemed necessary to estimate the prevalence of a previous condition or 30-day risk of an outcome affecting 10% of the study population with sufficient precision. Figure 1 presents the selection process of the databases for this study.

**Figure 1.**
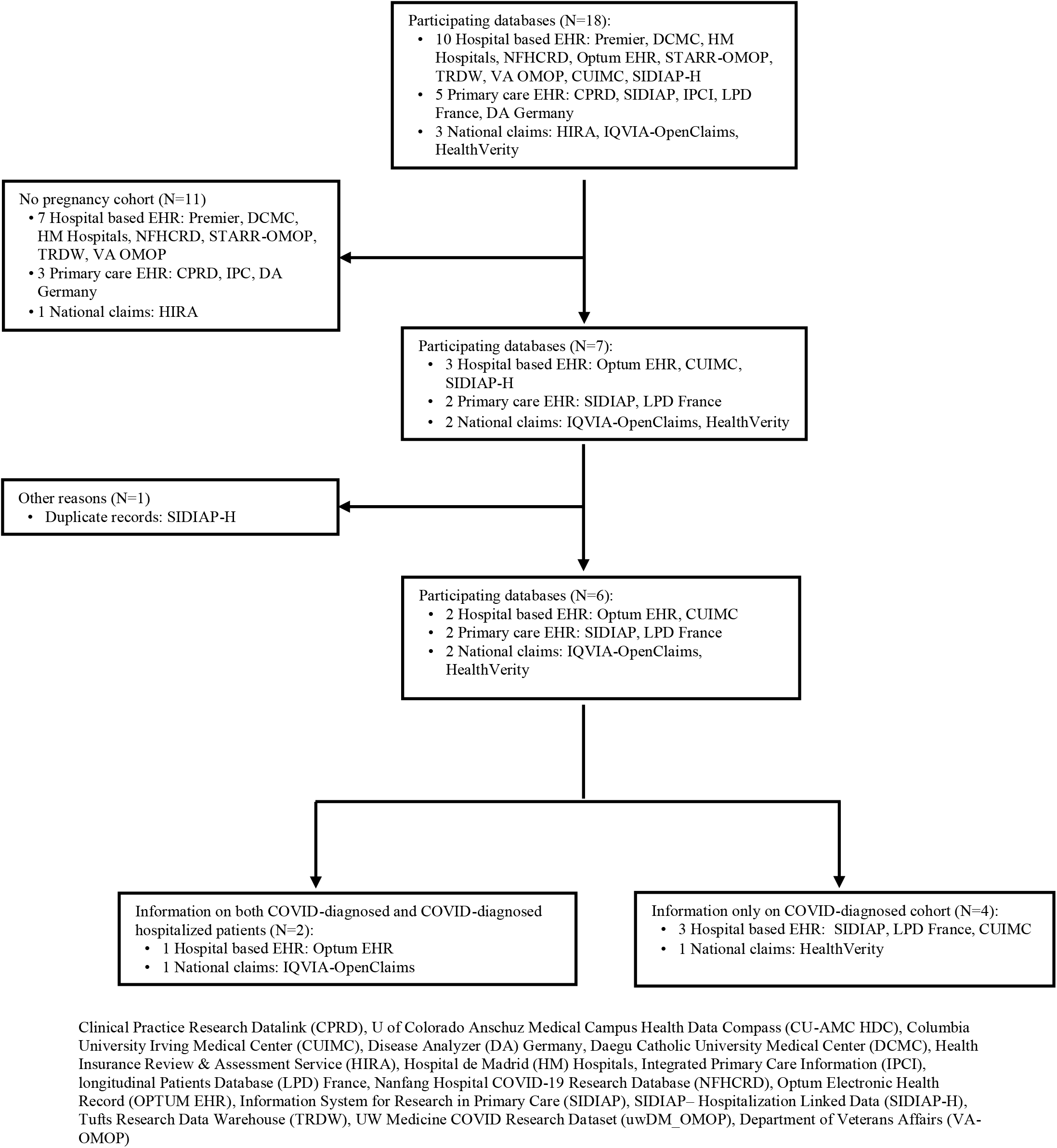
Database selection process flowchart.

Overall, six databases satisfied the inclusion criteria. United States (US) data included hospital EHR data from Columbia University Irving Medical Center (CUIMC), Optum® de-identified COVID-19 Electronic Health Record dataset (Optum) and national claims data from IQVIA-Open Claims and HealthVerity (HV). Spanish data came from the Information System for Research in Primary Care (SIDIAP). IQVIA Longitudinal Patients Database France (LPDFr) contributed data for French participants. A detailed description of the included data sources is available in Supplementary Table 1.

All reported data were extracted from the CHARYBDIS repository on 14^th^ September 2020.

### Study Participants

Two non-mutually exclusive COVID-19 cohorts were included in the study: 1) pregnant women with a COVID-19 diagnosis or a SARS-CoV-2 positive test between January 2020 and June 2020 (index date was the first earliest of the two events), referred to as *diagnosed cohort* (Supplementary Table 2) and, 2) pregnant women hospitalized between January 2020 and June 2020 with a COVID-19 diagnosis or a SARS-CoV-2 positive test 21 days before or after hospitalization date (index date), referred to as *hospitalized cohort*. All study participants were required to have at least 365 days of prior observation before index date to allow comprehensive capture of comorbidities.

Study participants could contribute information to both cohorts: pregnant women diagnosed with COVID-19 could be also included in the hospitalized cohort if COVID-19 was diagnosed during or at the time of hospitalization, or if they were hospitalized within 21 days after diagnosis. Similar comparison cohorts of pregnant women with seasonal influenza diagnosis or positive test in 2017-2018 were also included. The full description of the logic used for the *influenza diagnosed* cohort is provided at https://atlas.ohdsi.org/#/cohortdefinition/211.

Pregnancies were identified across all data sources using the same algorithm. Women aged between 12 and 55 years with pregnancy-related conditions, procedures or observations were identified in each data source. Study pregnancy period start date was set as the earliest date of occurrence of any pregnancy-related conditions, procedures or observations. Women who experienced any pregnancy-related events, delivery, or spontaneous abortion up to 300 days after pregnancy start date and not up to 60 days before the start of pregnancy were considered pregnant. Study pregnancy end date was the date of pregnancy-related events, delivery, or spontaneous abortion that occurred up to 300 days after the pregnancy start date. A detailed description of the pregnancy cohort and pregnancy events can be found <u>HERE</u>.

All women who were pregnant (as defined in the previous paragraph) at index date (as defined above) were eligible for inclusion in the study. Cohort participants were followed from their index date to the earliest of death, end of the observation period (June 2020), or 30 days after the index date.

### Baseline characteristics, medication use, and outcomes of interest

Baseline information on age, symptoms at index date, pregnancy related conditions up to 30 days prior and conditions up to 365 days before index date were identified. Conditions were ascertained based on the SNOMED CT hierarchy, with all descendant codes included. Detailed definitions of each condition can be found <u>HERE</u>.

All medications prescribed/dispensed during follow up and in the year prior were ascertained for characterization. Episodes of medication use were created to give the time span an individual is assumed to be exposed to an ingredient. Each drug episode started on the date of first drug exposure and ended on the observed end date if available, or were inferred (for example, based on the number of days of supply). Two prescriptions for the same drug with less than a 30-day gap were considered as part of the same drug episode. Individual medications were categorized using the Anatomical Therapeutic Chemical (ATC) classification. For the study of medications used for COVID-19, we assessed all medications included in at least two randomized controlled trials according to the COVID-19 clinical trial tracker^12^.

Study outcomes were categorized into two groups: 1) maternal complications and 2) pregnancy related outcomes. Maternal complications reported here included hospitalization, death, pneumonia, sepsis, acute respiratory disease syndrome (ARDS), chest pain/angina, cardiac arrhythmia, acute kidney injury (AKI), venous thromboembolic events (VTE), which includes pulmonary embolism and deep vein thrombosis, and cardiovascular (CV) events (including acute myocardial infarction, sudden cardiac death, ischemic stroke, intracranial bleed [hemorrhagic stroke] and heart failure). Pregnancy related outcomes included stillbirth, spontaneous abortion (miscarriage), livebirth, preterm delivery and caesarean section. The definition of each outcome is provided <u>HERE</u>.

### Statistical analyses

Demographics, comorbidities, and outcomes in each database were summarized as proportions, calculated by dividing the number of individuals within a given category by the total number of individuals. The proportion of individuals on each medication was determined as the percentage of subjects who had ≥1 day during the 30-day post-index period overlapping with a drug use episode for each medication or drug class of interest.

For databases with available data on both COVID-19 diagnosed and hospitalized cohorts, the distribution of symptoms, comorbidities, medications and outcomes in the diagnosed cohort (X axis) were compared to the hospitalized cohort (Y axis) in a scatter plot, with dots on the top-left quadrant of the plot indicating a higher prevalence among women hospitalized with COVID-19, and dots to the bottom-right of the diagonal line indicating a higher prevalence among all those diagnosed with COVID-19 as detailed above. Standardized mean differences (SMD) were calculated for the two cohorts, with an SMD >0.1 indicating a meaningful difference in the prevalence of a given covariate^13^. Covariates with SMD>0.1 were labelled in the scatter plot. Similarly, the distribution of conditions a year prior to index date, pregnancy-related conditions during the month prior to index date, symptoms at index date, and conditions and medications up to 30 days post-index date in the COVID-19 diagnosed cohort were compared to the influenza diagnosed cohort in a scatterplot.

Data were analyzed using a common analysis code that was developed for the OHDSI Methods library^10^. The code was run locally in each database (available at https://github.com/ohdsi-studies/Covid19CharacterizationCharybdis)^11^, and only aggregate results from each database were publicly shared in a dedicated interactive website: <u>CHARYBDIS</u>.

This study is descriptive in nature, and no causal inference is intended. Multivariable regression or adjustment for confounding was therefore considered out of remit, and not included in our study protocol.

All the data partners obtained Institutional Review Board (IRB) approval or exemption to conduct this study.

## RESULTS

A total of 8,598 pregnant women with COVID-19 were included in the diagnosed cohort (168 from France, 660 from Spain, and the remaining 7,770 from US databases, see Table 1), and 2,031 in the COVID-19 hospitalized cohort. An additional 12 data sources were screened from Germany, Netherlands, the United Kingdom, and South Korea, which could not contribute due to insufficient sample size (a flowchart of the database selection process is presented in Figure 1).

**Table 1.**
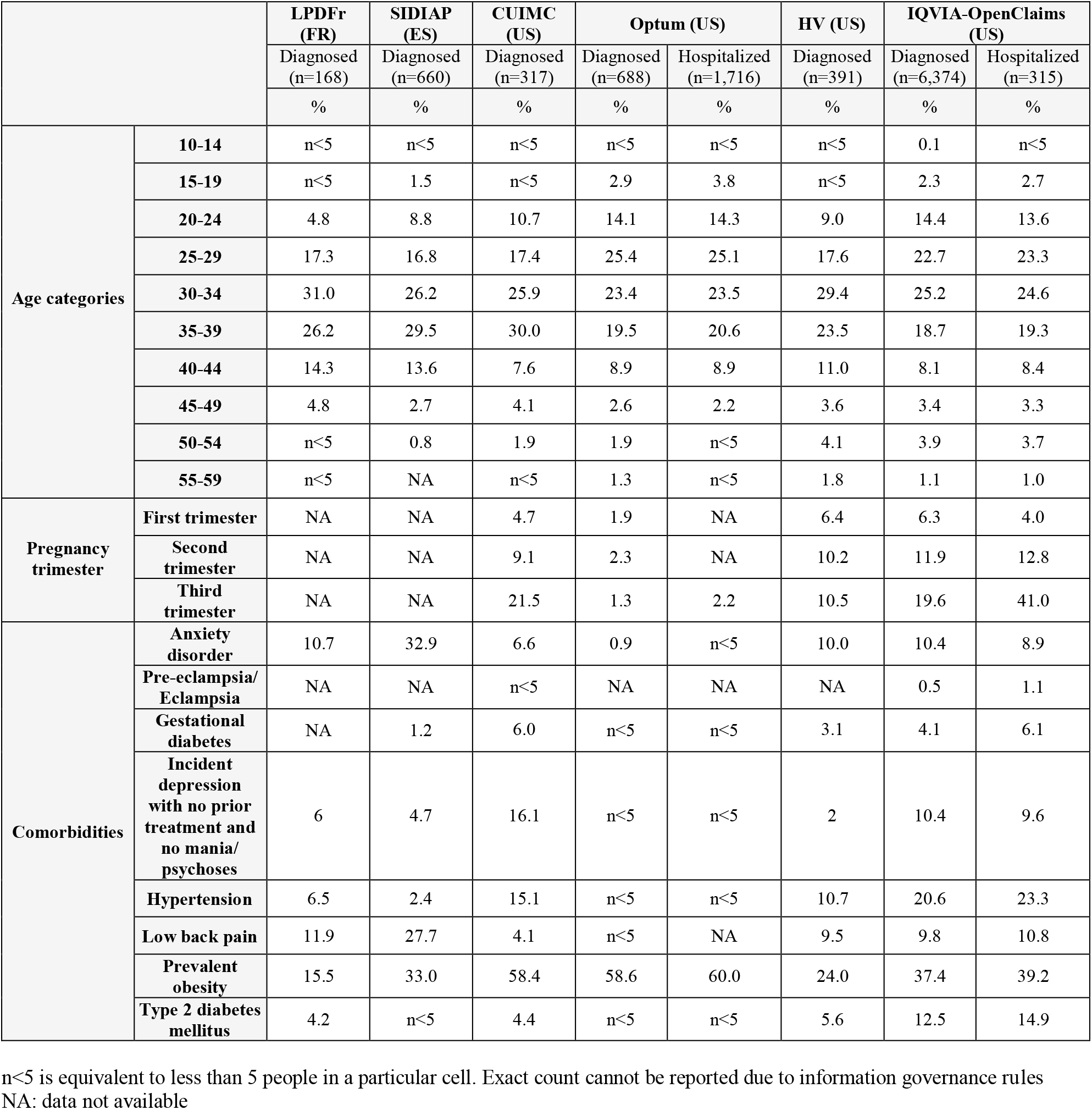
Baseline characteristics of study participants diagnosed or hospitalized with COVID-19, at time of diagnosis or hospitalization, respectively

|  |  | **LPDFr (FR)** | **SIDIAP (ES)** | **CUIMC (US)** | **Optum (US)** |  | **HV (US)** | **IQVIA-OpenClaims (US)** |  |
| --- | --- | --- | --- | --- | --- | --- | --- | --- | --- |
|  |  | Diagnosed (n=168) | Diagnosed (n=660) | Diagnosed (n=317) | Diagnosed (n=688) | Hospitalized (n=1,716) | Diagnosed (n=391) | Diagnosed (n=6,374) | Hospitalized (n=315) |
|  |  | % | % | % | % | % | % | % | % |
| **Age categories** | **10-14** | n<5 | n<5 | n<5 | n<5 | n<5 | n<5 | 0.1 | n<5 |
|  | **15-19** | n<5 | 1.5 | n<5 | 2.9 | 3.8 | n<5 | 2.3 | 2.7 |
|  | **20-24** | 4.8 | 8.8 | 10.7 | 14.1 | 14.3 | 9.0 | 14.4 | 13.6 |
|  | **25-29** | 17.3 | 16.8 | 17.4 | 25.4 | 25.1 | 17.6 | 22.7 | 23.3 |
|  | **30-34** | 31.0 | 26.2 | 25.9 | 23.4 | 23.5 | 29.4 | 25.2 | 24.6 |
|  | **35-39** | 26.2 | 29.5 | 30.0 | 19.5 | 20.6 | 23.5 | 18.7 | 19.3 |
|  | **40-44** | 14.3 | 13.6 | 7.6 | 8.9 | 8.9 | 11.0 | 8.1 | 8.4 |
|  | **45-49** | 4.8 | 2.7 | 4.1 | 2.6 | 2.2 | 3.6 | 3.4 | 3.3 |
|  | **50-54** | n<5 | 0.8 | 1.9 | 1.9 | n<5 | 4.1 | 3.9 | 3.7 |
|  | **55-59** | n<5 | NA | n<5 | 1.3 | n<5 | 1.8 | 1.1 | 1.0 |
| **Pregnancy trimester** | **First trimester** | NA | NA | 4.7 | 1.9 | NA | 6.4 | 6.3 | 4.0 |
|  | **Second trimester** | NA | NA | 9.1 | 2.3 | NA | 10.2 | 11.9 | 12.8 |
|  | **Third trimester** | NA | NA | 21.5 | 1.3 | 2.2 | 10.5 | 19.6 | 41.0 |
| **Comorbidities** | **Anxiety disorder** | 10.7 | 32.9 | 6.6 | 0.9 | n<5 | 10.0 | 10.4 | 8.9 |
|  | **Pre-eclampsia/Eclampsia** | NA | NA | n<5 | NA | NA | NA | 0.5 | 1.1 |
|  | **Gestational diabetes** | NA | 1.2 | 6.0 | n<5 | n<5 | 3.1 | 4.1 | 6.1 |
|  | **Incident depression with no prior treatment and no mania/psychoses** | 6 | 4.7 | 16.1 | n<5 | n<5 | 2 | 10.4 | 9.6 |
|  | **Hypertension** | 6.5 | 2.4 | 15.1 | n<5 | n<5 | 10.7 | 20.6 | 23.3 |
|  | **Low back pain** | 11.9 | 27.7 | 4.1 | n<5 | NA | 9.5 | 9.8 | 10.8 |
|  | **Prevalent obesity** | 15.5 | 33.0 | 58.4 | 58.6 | 60.0 | 24.0 | 37.4 | 39.2 |
|  | **Type 2 diabetes mellitus** | 4.2 | n<5 | 4.4 | n<5 | n<5 | 5.6 | 12.5 | 14.9 |

n<5 is equivalent to less than 5 people in a particular cell. Exact count cannot be reported due to information governance rules NA: data not available

Baseline characteristics for the study participants are reported in Table 1. Most pregnant women diagnosed with COVID-19 were 20 to 39 years old. France and Spain reported high proportions of pregnant women aged 40 years or older at the time of COVID-19 diagnosis (19.1% and 17.1%, respectively). Similar age distributions were seen for those hospitalized with COVID-19, e.g. 13.6% vs 14.3% aged 20-24, 23.3% vs 25.1% aged 25-29, 24.6% vs 23.5% aged 30-34, and 19.3% vs 20.6% aged 35-39 in IQVIA-OpenClaims and Optum respectively. Information on trimester of pregnancy at index date was available for a proportion of US participants (2.2% [hospitalized cohort in Optum] to 57.8% [hospitalized cohort in IQVIA-OpenClaims]). In the COVID-19 diagnosed cohort, diagnosis occurred most frequently in the third trimester of pregnancy (ranging from 1.3% in Optum to 21.5% in CUIMC). Similarly, COVID-19 was most commonly diagnosed in the third trimester of pregnancy for the COVID-19 hospitalized cohort (41.0% in IQVIA-OpenClaims).

The most common baseline comorbidities among both the COVID-19 diagnosed and hospitalized cohorts were obesity (e.g. 58.6% [Optum], 58.4% [CUIMC], 39.2% [IQVIA-OpenClaims]), anxiety (e.g. 32.9% [SIDIAP], 10.7% [LPDFr], 10.4% [IQVIA-OpenClaims]) and back pain (e.g. 27.7% [SIDIAP], 11.9% [LPDFr], 9.8% [IQVIA-OpenClaims]). As for the COVID-19 hospitalized cohort, additional conditions of interest based on IQVIA-OpenClaims included: hypertension (23.3%), type 2 diabetes mellitus (14.9%), gestational diabetes (6.1%) and pre-eclampsia/eclampsia (1.1%). When both COVID-19 diagnosed and hospitalized cohorts were characterized (in IQVIA-OpenClaims), differences with SMD ≥0.10 were noted in the prevalence of renal impairment (0.022 versus 0.051), anemia (0.155 versus 0.213), complication occurring during pregnancy (0.115 versus 0.195) and fetal condition affecting obstetrical care of mother (0.172 versus 0.250), all more common in the COVID-19 hospitalized than the COVID-19 diagnosed cohort (Figure 2a). When comparing the COVID-19 diagnosed cohort with the influenza diagnosed cohort, differences with SMD ≥0.10 were noted for respiratory failure (0.049 versus 0.003), pneumonia (0.12 versus 0.027), hypertension (0.206 versus 0.105), anemia (0.155 versus 0.072), chest pain or angina (0.33 versus 0.239) and obesity (0.195 versus 0.111), all more common in the COVID-19 diagnosed cohort (Figure 2b).

**Figure 2a.**
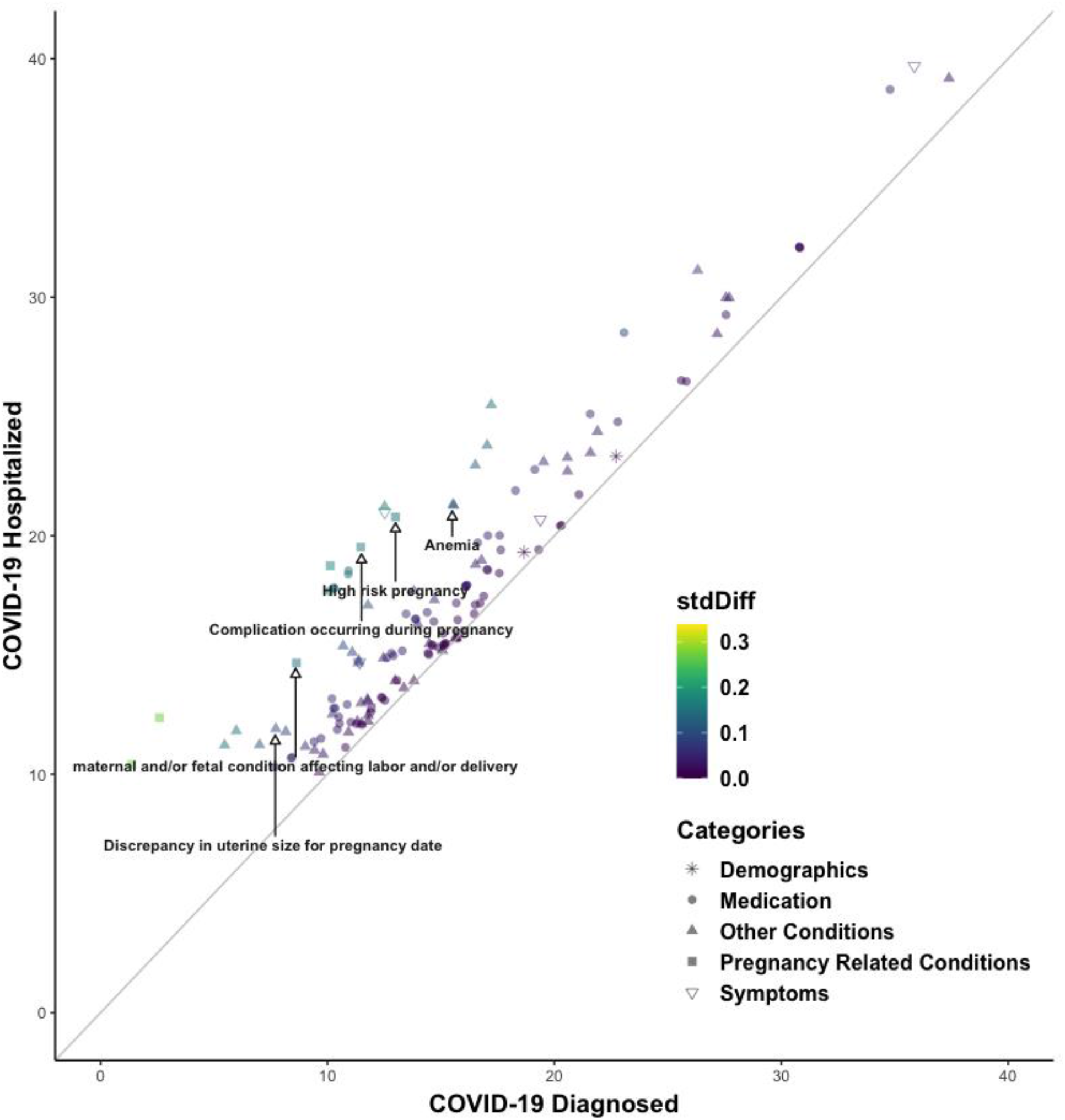
Scatter plot of prevalence of socio-demographics, medication use, comorbidities, symptoms and pregnancy outcomes in women diagnosed (X axis) versus hospitalized (Y axis) with COVID-19

**Figure 2b.**
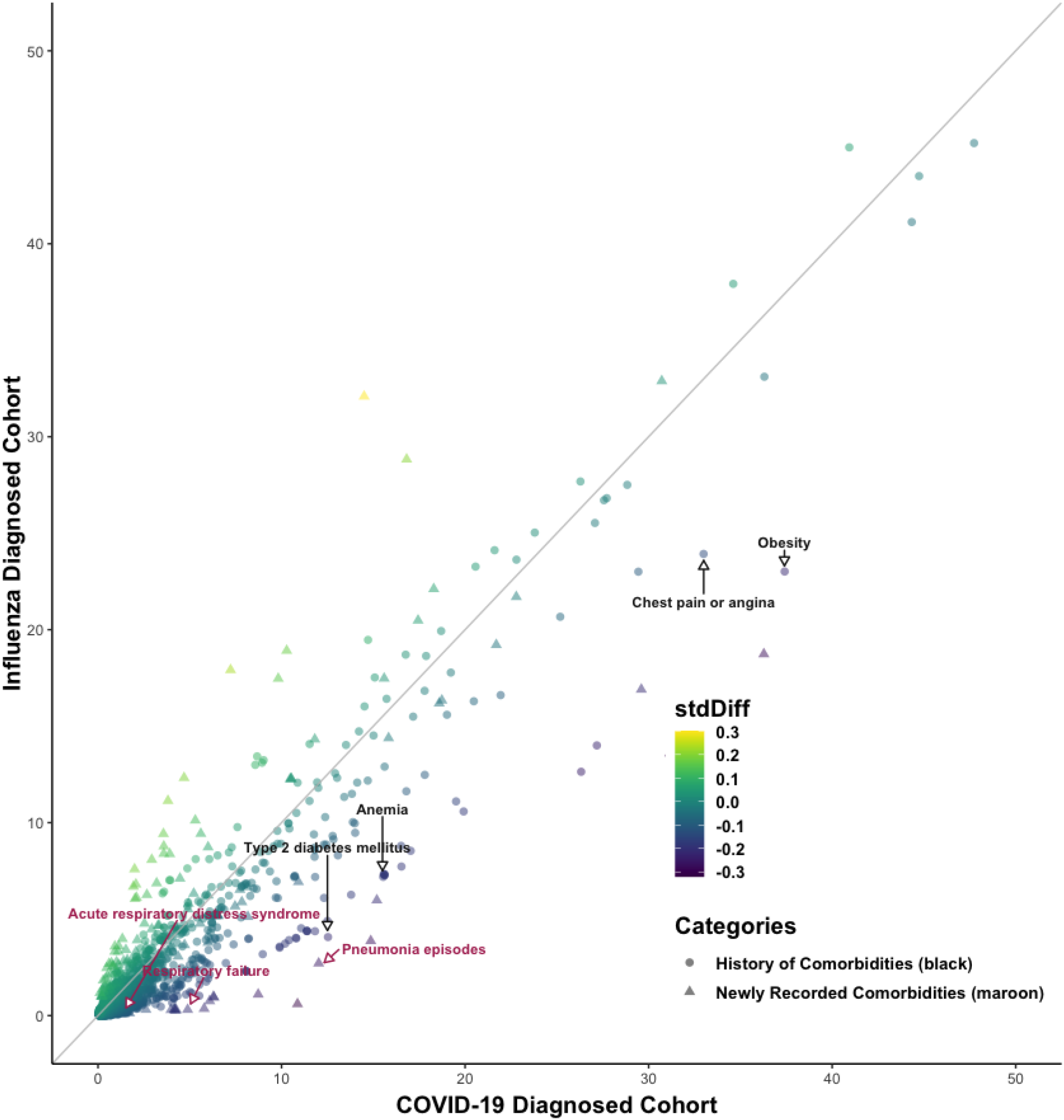
Scatter plot of prevalence of previous comorbidities and newly recorded comorbidities in pregnant women diagnosed with COVID-19 (X axis) versus diagnosed with influenza (Y axis)

Symptoms at index date were generally more common amongst pregnant women diagnosed with COVID-19 than those hospitalized with COVID-19 (in IQVIA-OpenClaims), including cough (9.3% vs. 4.1%) and fever (6.8% vs. 5.3%), respectively. Other symptoms such as dyspnea and malaise were similarly common in COVID-19 diagnosed and COVID-19 hospitalized patients (Figure 3a). When compared across all databases, the three most common symptoms amongst pregnant women diagnosed with COVID-19 were cough (from 1.2% [SIDIAP] to 13.3% [HV]), dyspnea (from 3.5% [CUIMC] to 8.0% [IQVIA-OpenClaims]), and fever (from 0.9% [SIDIAP] to 8.7% [HV]) respectively (Supplementary Figure 1a). Compared to symptoms seen in 27,510 women diagnosed with influenza, pregnant women diagnosed with COVID-19 (in IQVIA-OpenClaims) had a similar prevalence of cough, a lower prevalence of fever (6.8% in COVID-19 vs 10.8% in influenza), but a higher presence of dyspnea (8.0% vs 2.2%) and anosmia (0.7% vs 0.0%) (Figure 3b).

**Figure 3a.**
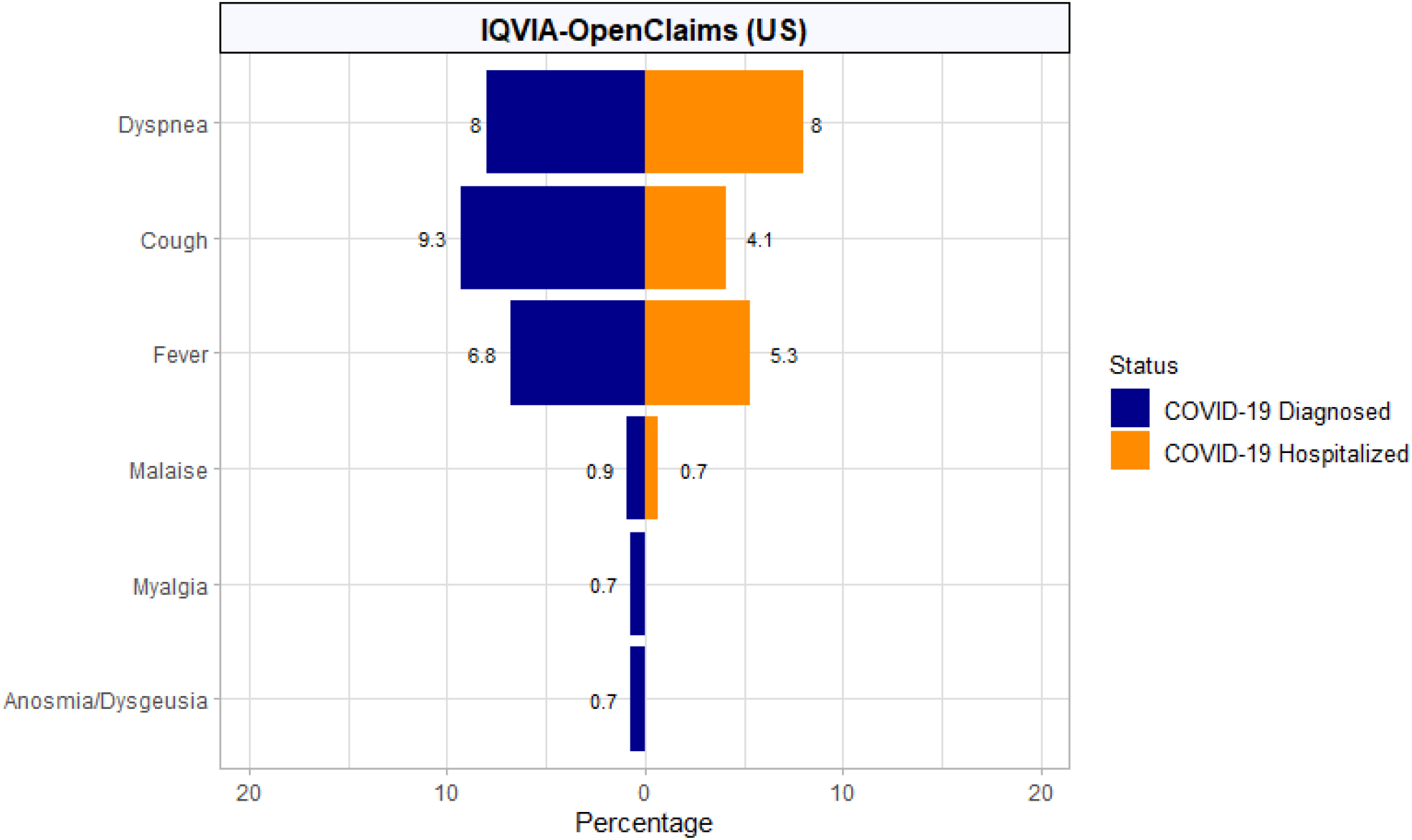
COVID-19 symptoms at index date amongst pregnant women diagnosed versus hospitalized with COVID-19.

**Figure 3b.**
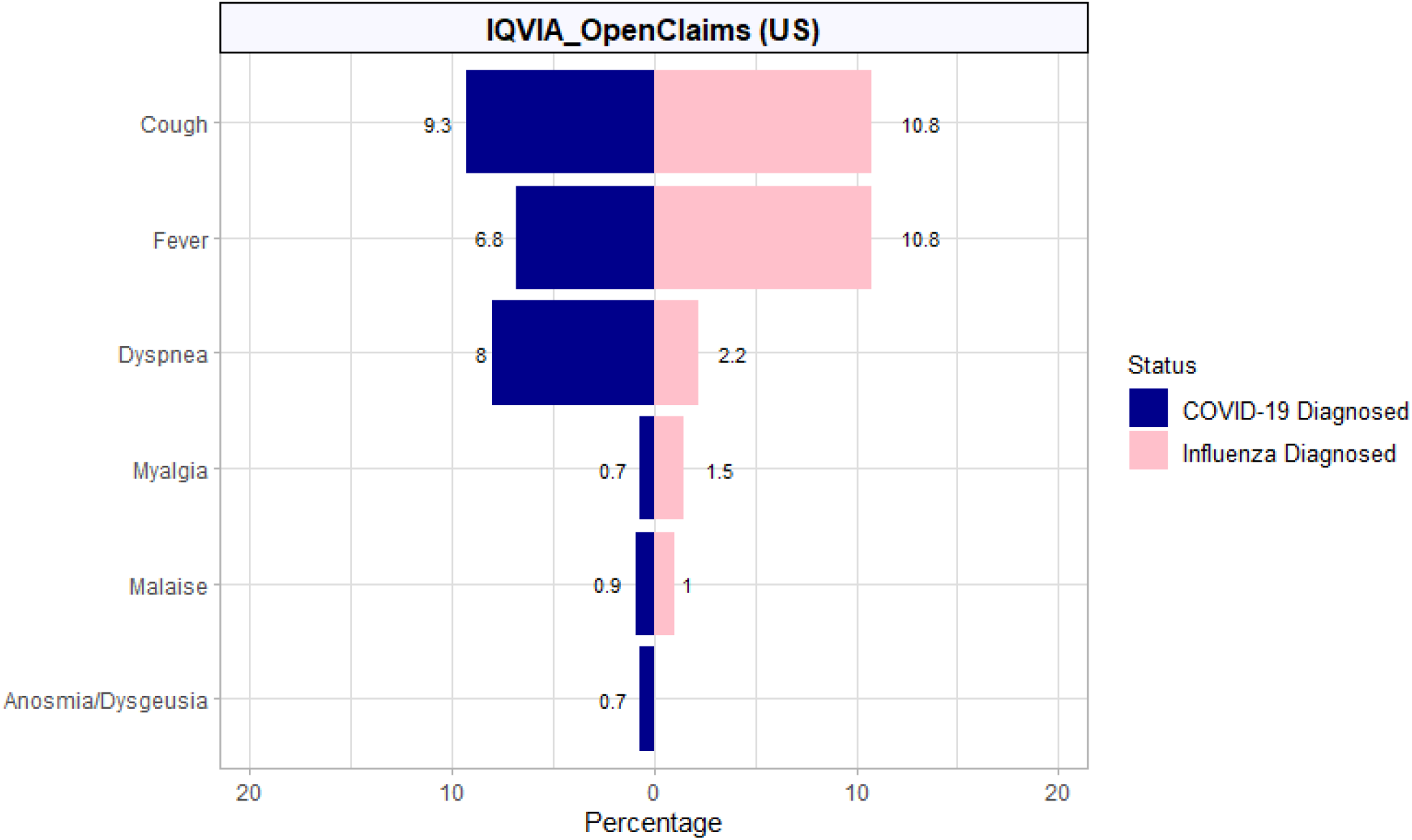
COVID-19 symptoms at index date amongst pregnant women diagnosed with COVID-19 versus diagnosed with influenza.

The use of medications during the 30 days following hospital admission with COVID-19 in pregnant women is depicted in Figure 4: split into repurposed/antiviral (Figure 4a) and adjunctive therapies (Figure 4b) obtained from Optum. In summary, the only two repurposed/antiviral therapies identified were azithromycin (18.1%) and hydroxychloroquine (5.4%). Conversely, the use of adjunctive therapies were common, including systemic corticosteroids (29.6%), antithrombotics (enoxaparin [24.0%,] heparin [15.8%] and aspirin [7.0%]), famotidine (20.9%), immunoglobulins (21.4%), and additional antibiotics (ceftriaxone [7.9%] and amoxicillin [3.5%]).

**Figure 4a.**
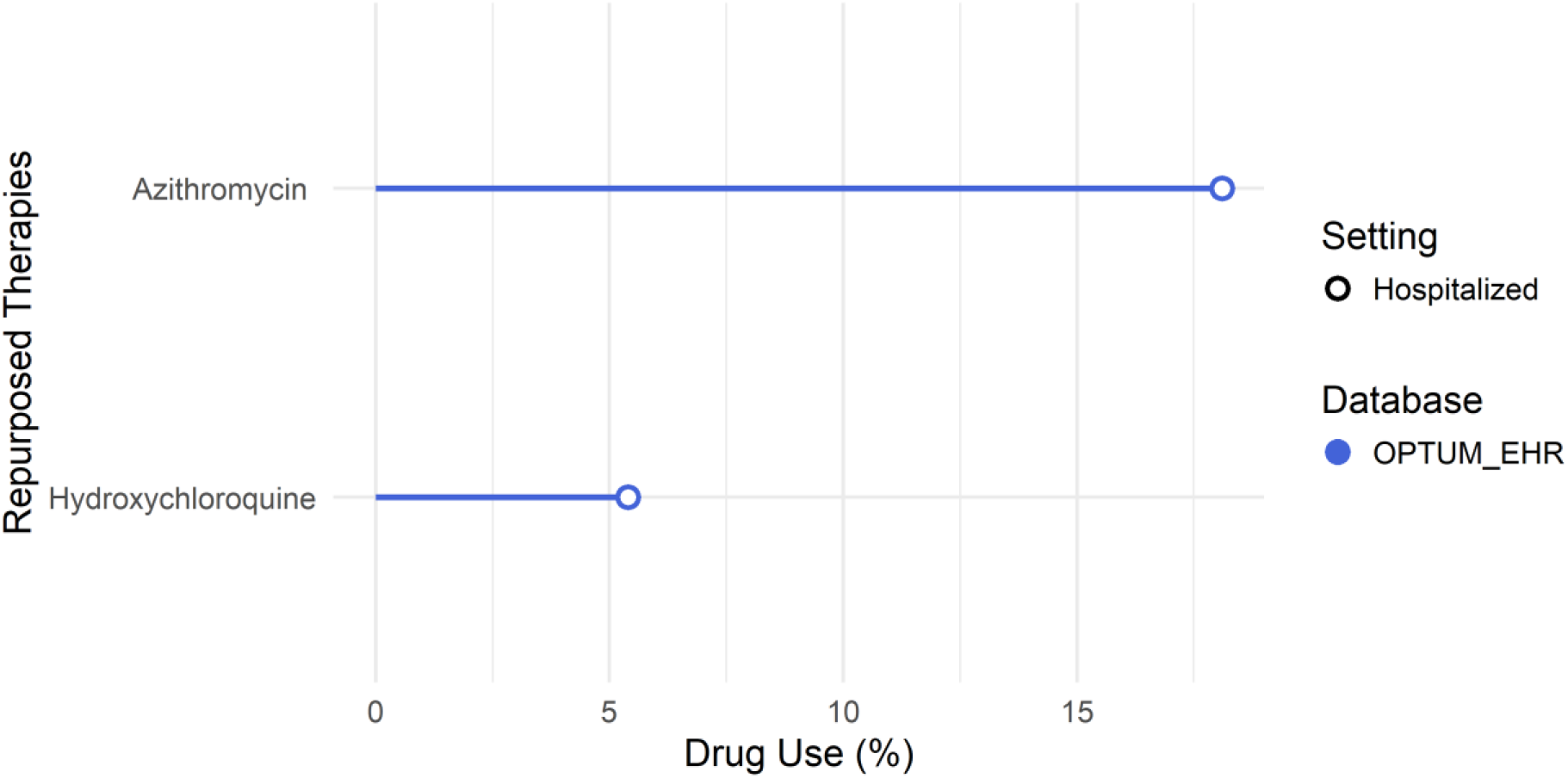
Repurposed therapies used during hospitalization/ inpatient treatment with COVID-19 in pregnant women.

**Figure 4b.**
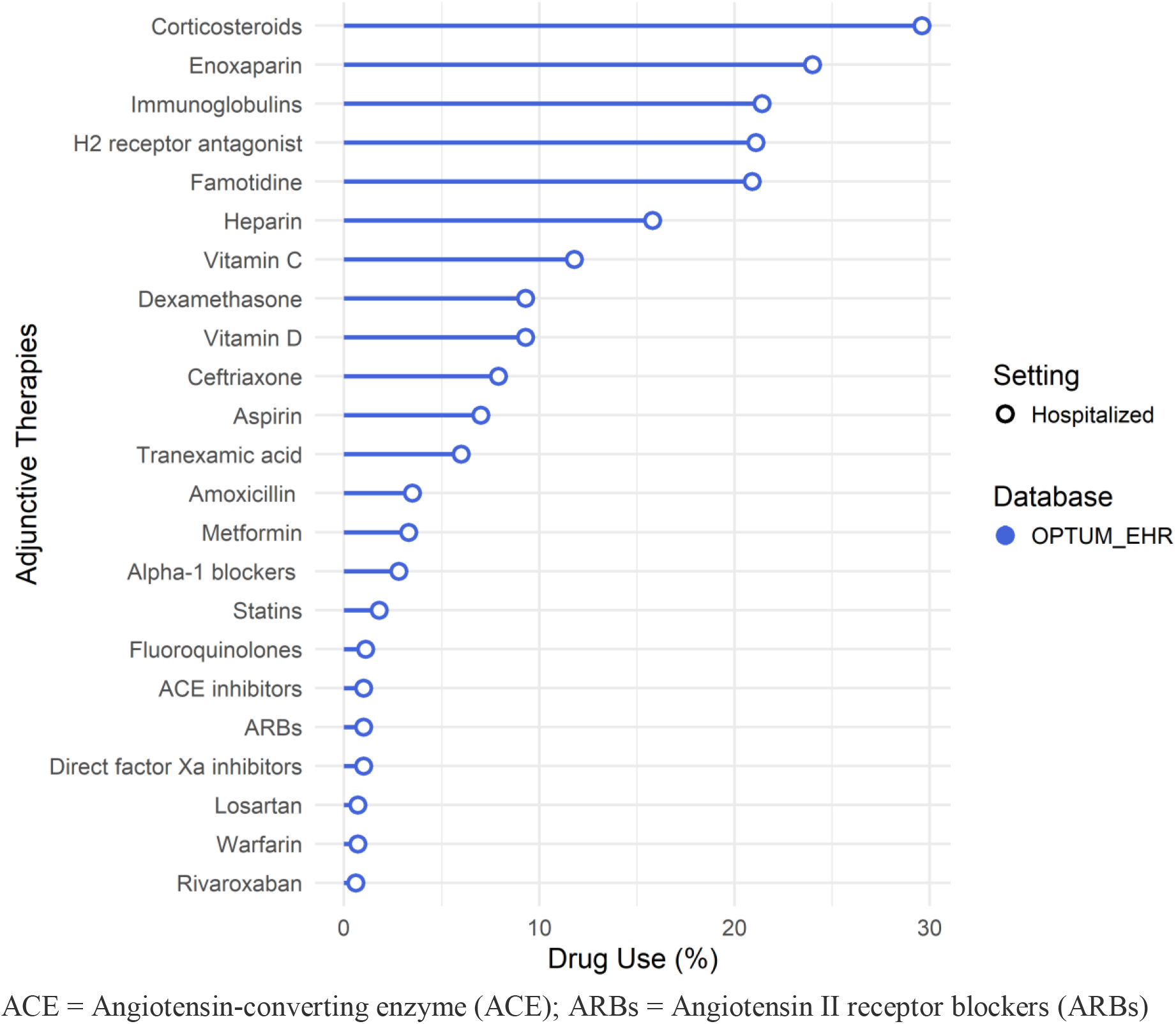
Adjunctive therapies used during hospitalization/ inpatient treatment with COVID-19 in pregnant women.

Pregnancy-related outcomes following the diagnosis of COVID-19 included (in order of frequency in IQVIA-OpenClaims) livebirth (15.8%), cesarean-section (4.4%), and preterm delivery (0.9%). The percentages of livebirth, cesarean-section and preterm delivery in the COVID-19 hospitalized cohort were higher (41.0%, 13.9% and 3.0% respectively). Spontaneous abortion or miscarriages were similar in both cohorts (1.6%). The percentage of stillbirth was 0.1% in the COVID-19 diagnosed cohort (Figure 5a).

**Figure 5a.**
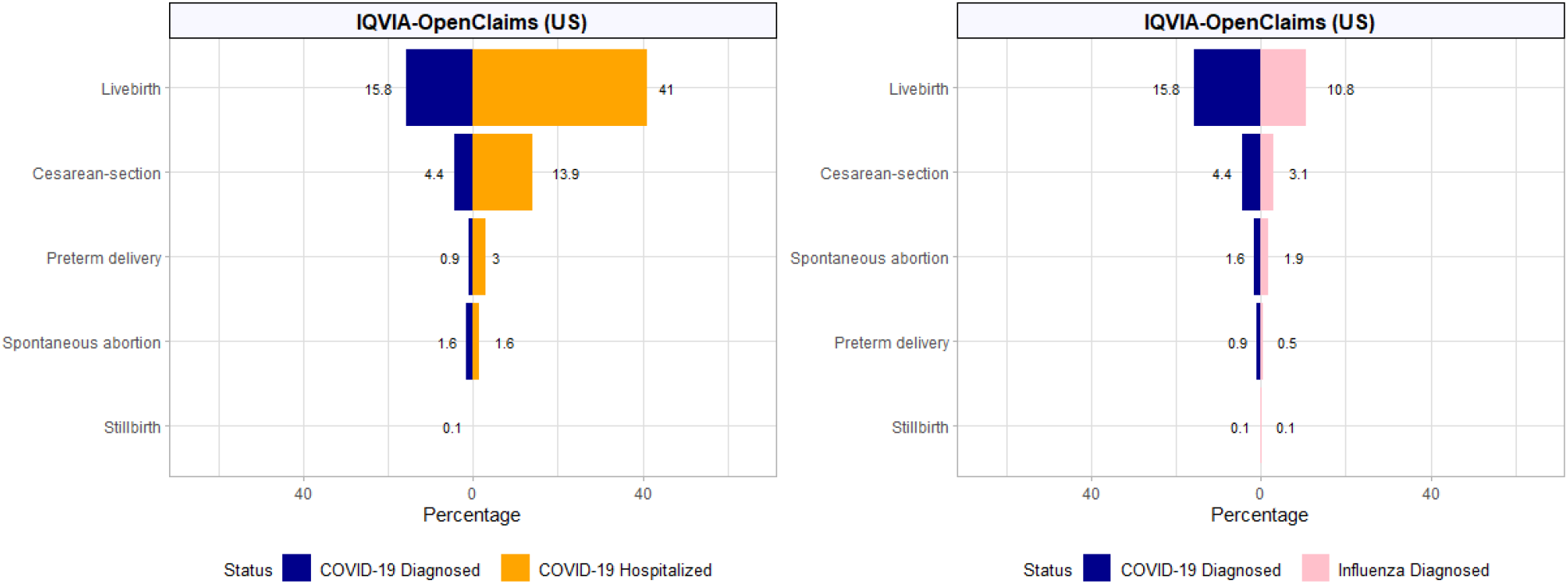
Pregnancy-related outcomes amongst women diagnosed versus hospitalized with COVID-19 (left) and diagnosed with COVID-19 versus diagnosed with influenza (right). Outcomes were within 30 days of diagnosis or hospitalization in the study cohorts, respectively.

Similar risks were observed in Optum (Supplementary Figure 2a). Compared to influenza, those diagnosed with COVID-19 in IQVIA-OpenClaims had a modestly higher proportions of livebirths (15.8% vs. 10.8%), cesarean-section (4.4% vs 3.1%), and preterm deliveries (0.9% vs 0.5%), but not of spontaneous abortion (1.6% vs 1.9%). The proportion of stillbirth was similar in both cohorts (0.1%) (Figure 5a).

Maternal complications in pregnant women diagnosed with COVID-19 (all from IQVIA-OpenClaims) included: pneumonia (12.0%), ARDS (4.0%), chest pain/angina (2.2%) and sepsis (2.1%). Less common yet relevant outcomes included cardiac arrhythmia (0.8%), AKI (0.6%), cardiovascular events (0.4%), heart failure (0.4%) and VTE (0.4%). All outcomes were more common in women hospitalized with COVID-19: pneumonia (22.8%), ARDS (12.6%), chest pain/angina (2.5%), sepsis (6.8%), cardiac arrhythmia (2.7%), chest pain/angina (2.5%), AKI (1.6%), cardiovascular events (1.3%), heart failure (1.2%) and VTE (1.1%) (Figure 5b). Fatality was negligible (less than 5 women died in CUIMC, and none in SIDIAP). The COVID-19 diagnosed cohort had a higher risk of 30-day maternal complications compared to pregnant women with seasonal influenza: hospitalization 29.6% vs 16.9%, pneumonia 12.0% vs 2.7%, ARDS 4.0% vs 0.3%, chest pain/angina 2.2% vs 1.8%, sepsis 2.1% vs 0.7%, cardiac arrhythmia 0.8% vs 0.2%, AKI 0.6% vs 0.1%, VTE 0.4% vs 0.1%, heart failure 0.4% vs 0.1%, and CV events 0.4% vs 0.1%, all in IQVIA-OpenClaims (Figure 5b).

**Figure 5b.**
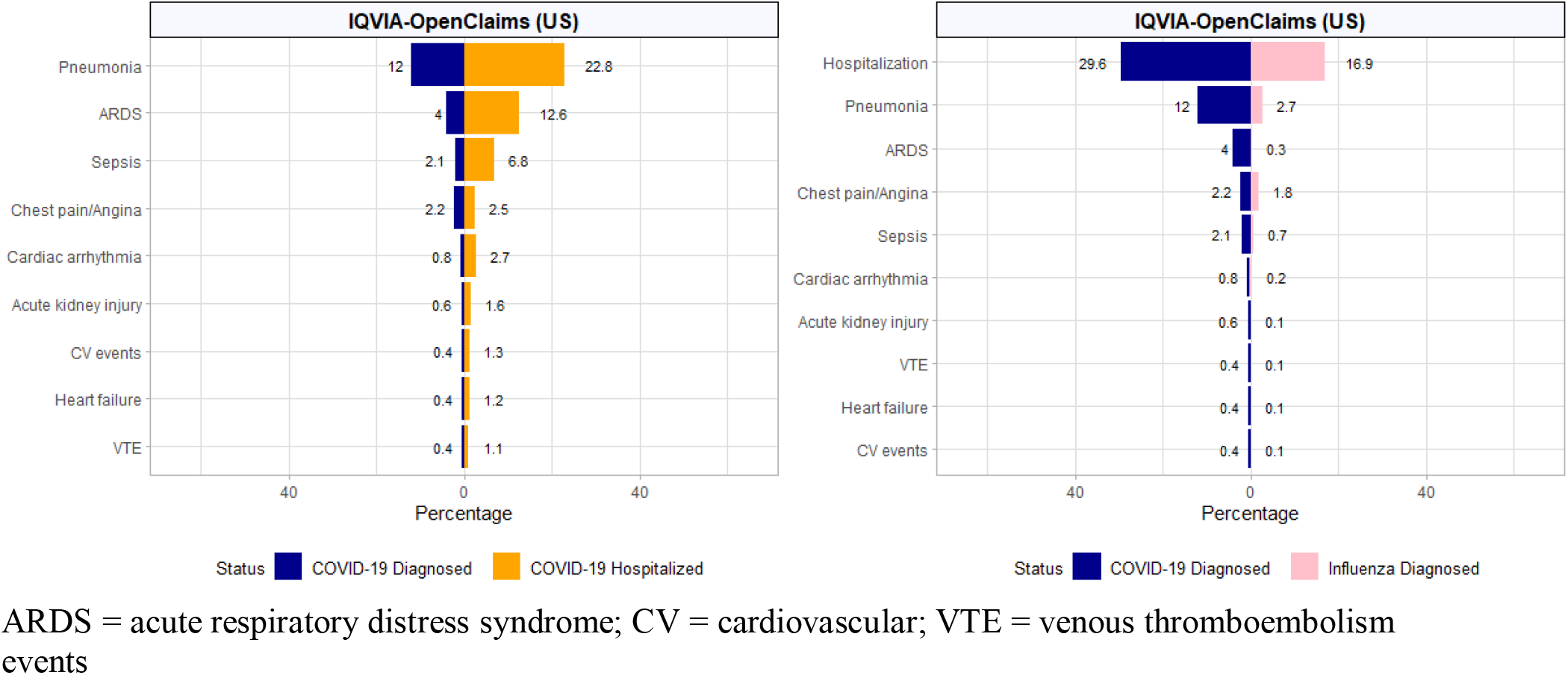
Maternal complications amongst women diagnosed versus hospitalized with COVID-19 (left) and diagnosed with COVID-19 versus diagnosed with influenza (right). Outcomes were within 30 days of diagnosis or hospitalization in the study cohorts, respectively.

## DISCUSSION

We present the largest study to date on pregnant women diagnosed and/or hospitalized with COVID-19, with linked baseline characteristics, symptoms, medication use, and outcomes from six databases across Europe and the US. Most pregnant women diagnosed with COVID-19 in the US were in the age range of 20-39 years old, while France and Spain reported a high proportion of women 40 or older. The pattern is consistent with trends from other published literature. One recent CDC study reported approximately one-fifth of pregnant women diagnosed with COVID-19 in the US were in the 35-44 age range^7^, while in France, approximately one-third of pregnant women diagnosed with COVID-19 were above 35^14^, and in Spain, approximately half of the pregnant women diagnosed with COVID-19 were above 35^15^. The average age of pregnant women in France and Spain have also been reported to be older than those in the US (26.4 [US], 28.5 [France] and 30.7 [Spain])^16^. We are however, unable to determine whether our finding is a reflection of the baseline age range difference during pregnancy between countries, or whether because these women were older and pregnant which placed them at greater risk of being infected with COVID-19.

Regarding previous comorbidities, obesity, hypertension, and diabetes were more prevalent in our hospitalized cohort, in line with previous literature^6^. Renal impairment and anemia were also more common in the COVID-19 hospitalized cohort, again indicating higher disease severity or worse outcomes than the COVID-19 diagnosed cohort. Anxiety was highly prevalent amongst pregnant women with COVID-19 in our study, which aligned well with another COVID-19 study, conducted in Canada by Lebel C et al^17^. The study reported that pregnant women suffered high rates of anxiety and depression during the pandemic, in part due to social isolation, concerns about availability of prenatal care, relationship strains, and concerns about threats of COVID-19 and associated therapies and outcomes to their baby^17^. Back pain was also common among our diagnosed COVID-19 cohort, which are usually occurring during normal pregnancy.

While there is heterogeneity in the presentation of symptoms across databases, there is a consistent appearance of cough, fever and dyspnea on index date in both the COVID-19 diagnosed and COVID-19 hospitalized cohorts. Our findings are consistent with previous studies^6, 7^. The frequency of reported symptoms in our study is generally lower than in other studies, suggesting an underestimation in the coding of symptoms in the form of structured data in busy actual care settings. It is also possible that many of those diagnosed (or hospitalized) may have been seen for pregnancy-related issues (e.g. prenatal visit, preterm delivery or contractions), with COVID-19 being diagnosed due to standard screening procedures, therefore increasing the proportion of asymptomatic diagnoses. We compared symptoms and outcomes of pregnant women diagnosed with COVID-19 with those diagnosed with influenza for benchmarking purposes. Cough, fever and myalgia were more commonly reported in pregnant women with influenza, which is not surprising, as these are well-known flu symptoms. One key finding in our study is that, while the percentages are small, symptoms such as dyspnea and anosmia were more common in COVID-19 diagnosed pregnant women than in pregnant women diagnosed with influenza, which aligns with what is known about COVID-19. This information is clinically relevant for differential diagnosis especially during the influenza season when COVID-19 and influenza may coexist. In addition, this information is also essential from an epidemiological perspective, as COVID-19 is known to remain contagious for a longer period of time after a positive test than influenza^18^.

There is a plethora of medications used in clinical practice among hospitalized pregnant women, mostly consisting of adjunctive therapies. Use of systemic corticosteroids was high, and could be partially attributable to pregnancy-related indications such as accelerating fetal lung development in pregnant women at risk of preterm delivery, or to maternal disease e.g. asthma exacerbations. In addition, nearly 1 in 4 hospitalized pregnant women were on antithrombotics, most likely as a prophylaxis against venous thromboembolism. Use of immunoglobulins was also high (nearly 1 in 5), although due to the nature of our study, we did not have the granularity in data to determine if they were anti-D immunoglobulin (used to neutralized Rhesus D positive antigens that may have entered the mother’s blood during pregnancy), or for the off–label use in the treatment of COVID-19. Use of famotidine was also high, possibility for the treatment of COVID-19^19, 20^, and also for the treatment of heartburn or gastroesophageal reflux disease in pregnancy, or for the prevention of gastrointestinal bleeding in hospitalized patients. Approximately 1 in 4 hospitalized pregnant women were also on azithromycin. The safety on the use of azithromycin in pregnancy is debatable as some studies have found a higher risk of birth defects^21^, while others have reported no increased risk^22-24^. While azithromycin may been prescribed for pneumonia, there is also a possibility that azithromycin was prescribed in combination with hydroxychloroquine, due to alleged antiviral efficacy against COVID-19. Despite the widespread use in the initial stage of the pandemic, we now know that this combination is not effective and potentially harmful^25^. Our study has added insight to prescribing patterns among pregnant women hospitalized with COVID-19, especially in the early months of the pandemic when it was not well understood which medications were truly effective. Our findings should however, be interpreted with caution as we were not able to determine the specific indication of these medications (i.e. prescribed for COVID-19 treatment or other pregnancy-related indication), or the stage of pregnancy at which these medications were administered.

In terms of 30-day pregnancy-related outcomes, our study findings were consistent with other published literature^6^. Overall, the proportion of cesarean-section and preterm births was higher in the hospitalized cohort than the diagnosed cohort, but this could in part be explained by hospital-based pre-scheduled cesarean-sections and preterm births leading to testing and hence COVID-19 diagnosis. This matches the baseline characteristics from this study, where the highest percentage of COVID-19 in the diagnosed and hospitalized cohorts were in the third trimester. The proportion of stillbirth was very low, consistent with a recently published systematic review^6^. Although rare in our study, women diagnosed with COVID-19 had slightly higher proportions of caesarean section and preterm births but similar proportions of stillbirth compared to pregnant women with influenza.

The proportion of various maternal complications varied across databases, though pneumonia and ARDS were consistently reported as the most common outcomes, seen in 2% to 12% and 1% to 4% of COVID-19 diagnosed women, respectively, across databases. While the magnitude of our findings was lower than that reported in previous studies, the most common maternal complications identified were identical^6^. Fatality was negligible (<5 women died in any of the participating databases), in line with a recent systematic review^6^. Age (mostly <40) and the modulating and anti-inflammatory actions of estradiol and progesterone have been proposed as the key reasons underlying the observed low COVID-19 fatality in pregnant women^26^. As expected, outcomes were worse amongst women diagnosed with COVID-19 who needed hospitalization: 1 in 4 developed pneumonia, 1 in 8 ARDS. When comparing pregnant women diagnosed with COVID-19 and with influenza, maternal outcomes were worse in the former, with almost double risk of hospitalization, a 6-fold higher risk of pneumonia, >10-fold higher risk of ARDS, 3-fold higher risk of sepsis, 6-fold higher risk of AKI, and a 4-fold higher risk of arrhythmia, heart failure, cardiovascular and VTE events. The trend of severity in our study suggests that COVID-19 infection has higher risk of adverse maternal outcomes than influenza, and is consistent with findings from a similar comparison based on the general population^27^.

### Strengths and Limitations

Our study has both strengths and weaknesses. Firstly, the use of routinely collected data (EHR and claims) consistently leads to an underestimation of absolute risks due to incomplete recording of symptoms and potentially some comorbidities^22^. Secondly, the lack of information on or misclassification of indication for a given therapy makes it difficult to differentiate medications used for the treatment of COVID-19 vs the treatment of pregnancy-related indication. Corticosteroids are an example of a prescription commonly observed in our data that could be the consequence of respiratory distress due to COVID-19 but also potentially for the acceleration of fetal lung development in women at risk of preterm delivery. Similarly, hospitalization is reported primarily as a consequence of COVID-19 diagnosis, but it is also likely to be concomitant with a COVID-19 diagnosis. For example, in the US, many centers began universal screening of all pregnant women presenting to the hospital regardless of reason (i.e. spontaneous labor, schedule cesareans, or evaluation of other pregnancy related condition) or presence of symptoms after early reports from New York cited asymptomatic COVID-19 positive testing rates of 14-66%^28, 29^. Finally, disparities in data sources due to different countries and healthcare settings may result in important differences in coding practice, making comparisons between databases difficult and increasing ranges in the observed frequencies for comorbidities, symptoms and outcomes. However, comparisons versus influenza were done in a single, very large database, which reinforces internal validity of these comparisons.

This study also has strengths. The use of a common data model, centrally developed programs and a distributed network strategy allowed us to consistently harmonize and analyze the largest dataset on pregnant women with COVID-19 to date, including almost 8,600 women from three countries in two continents. Such large sample size allows greater power to capture less common events, including organ failure (e.g., AKI) and VTE events. Another strength is the relative speed with which such a large study was completed, which is key in a rapidly evolving public health crisis. The use of routinely collected data allows for a realistic characterization of actual practice in busy clinical settings through the minimization of selection bias and Hawthorne effects^30^. In addition, the use of existing data is efficient, maximizes the value of readily available information and enables inclusion of wider populations compared to primary data collection studies that tend to focus on academic and specialized treatment^31^.

## CONCLUSION

In this large international cohort of pregnant women with COVID-19, previous renal impairment and anemia appeared associated with hospitalization. Anosmia and dyspnea were indicative symptoms of COVID-19, potentially helpful for the clinical diagnosis of COVID-19 in pregnant women. Regarding their management, we listed multiple medications used to treat inpatient pregnant women hospitalized with COVID-19, many of them with low evidence of antiviral efficacy. As more data becomes available, we hope to see a shift to use of treatments that are demonstrated to be safe and effective in pregnant women, a population vulnerable to adverse drug reactions. Finally, we report negligible fatality rates, but relatively common maternal complications in COVID-19 (e.g. pneumonia, ARDS and sepsis), much worse than following infection with influenza in pregnancy. More research is needed on the management of COVID-19 in pregnancy, including diagnosis and the management of more severe forms of the disease to maximize the benefit-risk of any proposed treatments and clinical outcomes.

## Data Availability

https://data.ohdsi.org/Covid19CharacterizationCharybdis/

## Ethical approval

All the data partners received Institutional Review Board (IRB) approval or exemption. The research was approved by the Columbia University Institutional Review Board as an OHDSI network study. The use of SIDIAP was approved by the Clinical Research Ethics Committee of the IDIAPJGol (project code: 20/070-PCV. The use of the HealthVerity and Optum databases was reviewed by the New England Institutional Review Board (IRB) and were determined to be exempt from broad IRB approval.

The use of IQVIA-OpenClaims and IQVIA LPD France were exempted from IRB approval.

## Transparency declaration

Lead authors affirm that the manuscript is an honest, accurate, and transparent account of the study being reported; that no important aspects of the study have been omitted; and that any discrepancies from the study as planned have been explained.

## Contributorship statement

KK, TDS, APU, PR, AS, TMA, AG, DPA, FN, DRM conceived and designed the study. SLD, TF, KEL, MEM, KN, JDP, CGR, NHS, PR, KK and TDS coordinated data contributions at their respective sites. AGS, TF, SFB, JDP, KK and TDS analyzed the data; AG and LL produced the figures and tables. LL, AG, WURA, KS, OA, HA, JJ, PC, CA, VS, LZ, RJC, AVM searched the literature. AS, MAS and PR developed the analysis tools. LL, AG, DPA wrote the first draft. All authors contributed to the interpretation of results, revision and development of the manuscript, and reviewed and approved the final version of the manuscript.

## Acknowledgements

We would like to acknowledge the patients who suffered from or died of this devastating disease, and their families and caretakers. We would also like to thank the healthcare professionals involved in the management of COVID-19 during these challenging times in all healthcare settings, from primary care to intensive care units, and all around the world.

## Data sharing statement

Analyses were performed locally in compliance with all applicable data privacy laws. Although the underlying data is not readily available to be shared, authors contributing to this paper have direct access to the data sources used in this study. All results (e.g. aggregate statistics, not presented at a patient-level with redactions for minimum cell count) are available for public inquiry. These results are inclusive of site-identifiers by contributing data sources to enable interrogation of each contributing site. All analytic code and result sets are made available at: https://github.com/ohdsi-studies/Covid19CharacterizationCharybdis

## Copyright/licence for publication

“The Corresponding Author has the right to grant on behalf of all authors and does grant on behalf of all authors, a worldwide licence to the Publishers and its licencees in perpetuity, in all forms, formats and media (whether known now or created in the future), to i) publish, reproduce, distribute, display and store the Contribution, ii) translate the Contribution into other languages, create adaptations, reprints, include within collections and create summaries, extracts and/or, abstracts of the Contribution, iii) create any other derivative work(s) based on the Contribution, iv) to exploit all subsidiary rights in the Contribution, v) the inclusion of electronic links from the Contribution to third party material where—ever it may be located; and, vi) licence any third party to do any or all of the above.”

