## Supplementary Materials for "“Clinical characteristics, symptoms, management and health outcomes in 8,598 pregnant women diagnosed with COVID-19 compared to 27,510 with seasonal influenza in France, Spain and the US: a network cohort analysis”"

**Supplementary Table 1. Description of databases included in the study**

| Country | Name | Description |
|---|---|---|
| US | HealthVerity | This HealthVerity derived data set contains de-identified patient information with an antibody and/or diagnostic test for COVID-19 linked to all available Medical Claims and Pharmacy Data from select private data providers participating in the HealthVerity marketplace. |
| US | Optum EHR | Optum's Electronic Health Record data, a medical records database for patients receiving a COVID-19 diagnosis record or lab test for SARS-CoV-2. |
| Spain | Information System for Research in Primary Care (SIDIAP) | The Information System for Research in Primary Care (SIDIAP; www.sidiap.org) is a primary care records database that covers approximately 7 million people, equivalent to an 80% of the population of Catalonia, North-East Spain. Healthcare is universal and tax-payer funded in the region, and primary care physicians are gatekeepers for all care and responsible for repeat prescriptions. |
| France | LPD | Computerised network of physicians including GPs who contribute to a centralised database of anonymised patient EMR. Currently, >1200 GPs from 400 practices are contributing to the database covering 7.8M patients in France. |
| US | IQVIA Open Claims | Pre-adjudicated claims covering over 300 Million lives (~80% of the US) collected from office-based physicians and specialists via office management software and clearinghouse switch sources for the purpose of reimbursement. |
| US | Columbia University Irving Medical Center | The clinical data warehouse of NewYork-Presbyterian Hospital/Columbia University Irving Medical Center, New York, NY, based on its current and previous electronic health record systems, with data spanning over 30 years and including over 6 million patients |

**Supplementary Table 2. Definitions and codes used to identify persons with a COVID-19 diagnosis record or a SARS-CoV-2 positive test with at least 365d prior observation**

The below tables summarize the concepts used to identify patients tested for SARS-CoV-2 and patients tested positive for SARS-CoV2. The full description of the logic used is provided at https://atlas.ohdsi.org/#/cohortdefinition/200 for *diagnosed* and https://atlas.ohdsi.org/#/cohortdefinition/197 for *hospitalized* cohorts.

**COVID-19 conditions**

| Id | Name | Vocabulary |
|---|---|---|
| 439676 | Coronavirus infection | SNOMED |
| 4100065 | Disease due to Coronaviridae | SNOMED |
| 37311060 | Suspected disease caused by 2019-nCoV | SNOMED |
| 37311061 | Disease caused by 2019-nCoV | SNOMED |

**COVID-19 specific test positive**

| Id | Name | Vocabulary |
|---|---|---|
| 37310282 | 2019 novel coronavirus detected | SNOMED |
| 37310281 | 2019 novel coronavirus not detected* | SNOMED |
| 756055 | Measurement of Severe acute respiratory syndrome coronavirus 2 (SARS-CoV-2) | OMOP Extension |

*used to only include measurements have a value equal detected, positive or present

**Supplementary Figure 1a. COVID-19 symptoms at index date amongst pregnant women diagnosed versus hospitalized with COVID-19 across all databases**

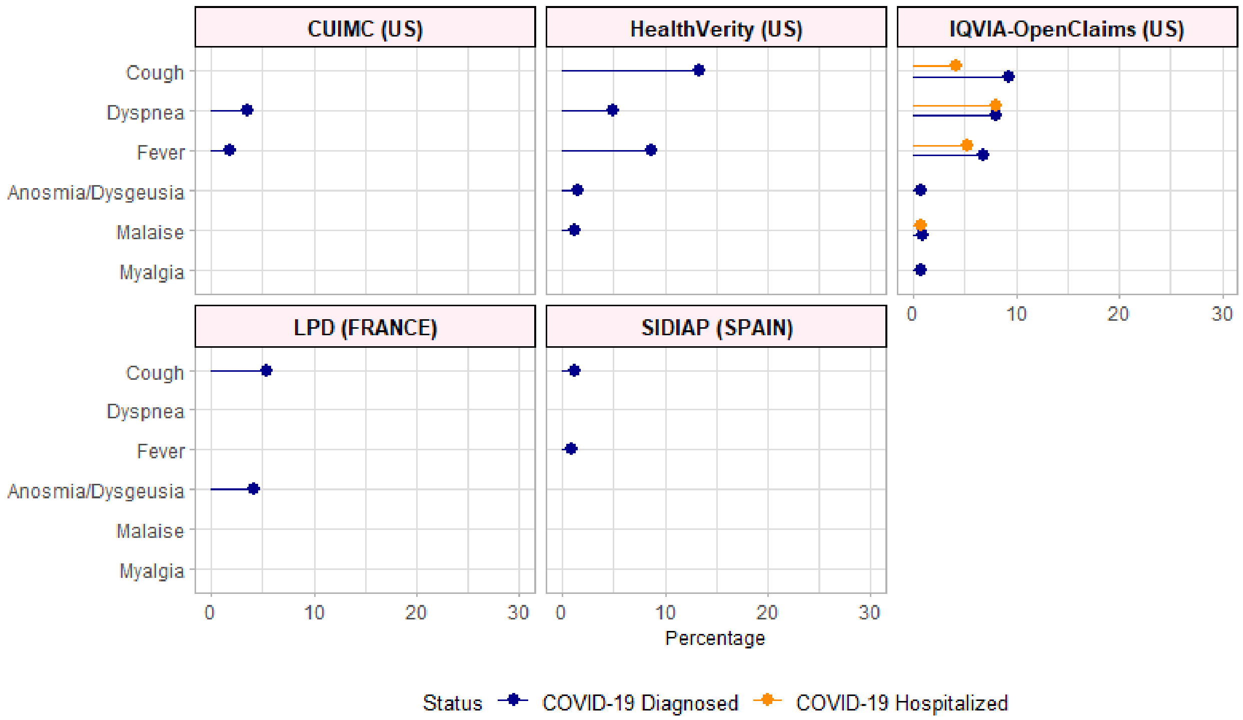


**Supplementary Figure 1b. COVID-19 symptoms at index date amongst pregnant women diagnosed with COVID-19 versus diagnosed with seasonal influenza across all databases**

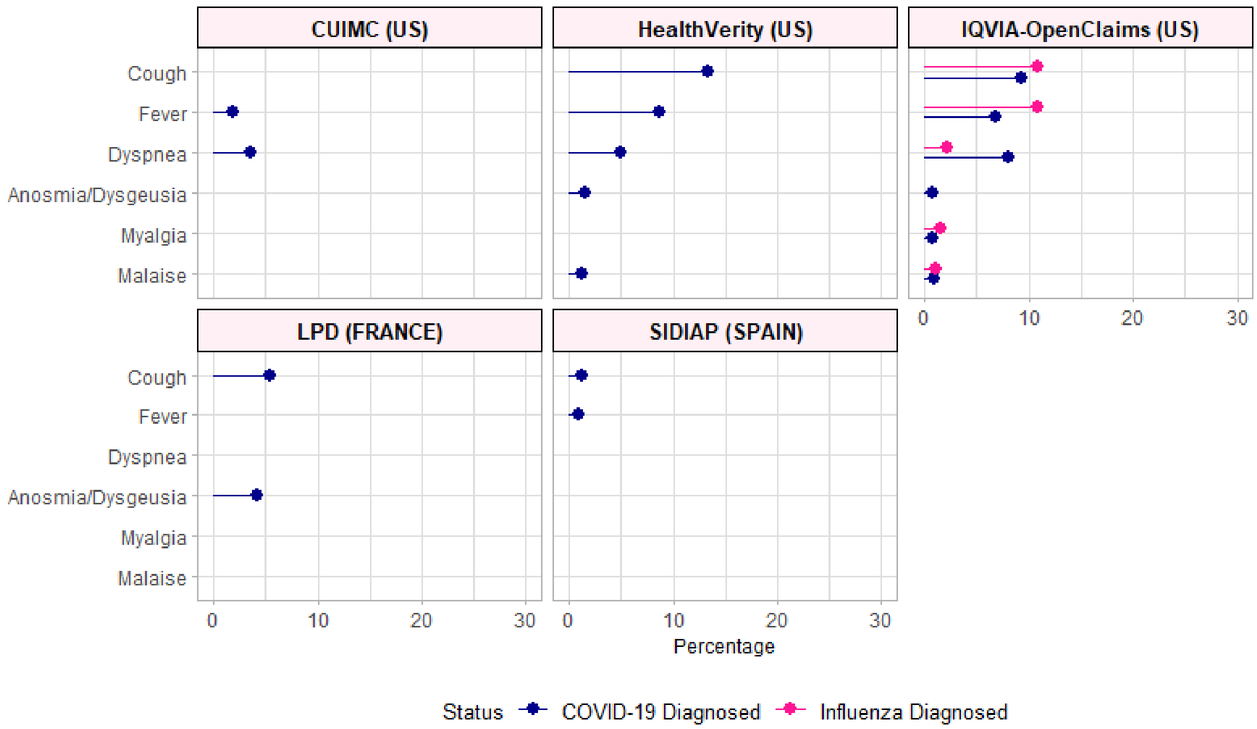

**Supplementary Figure 2a. Maternal complications and pregnancy-related outcomes amongst women diagnosed versus hospitalized with COVID-19 across all databases**

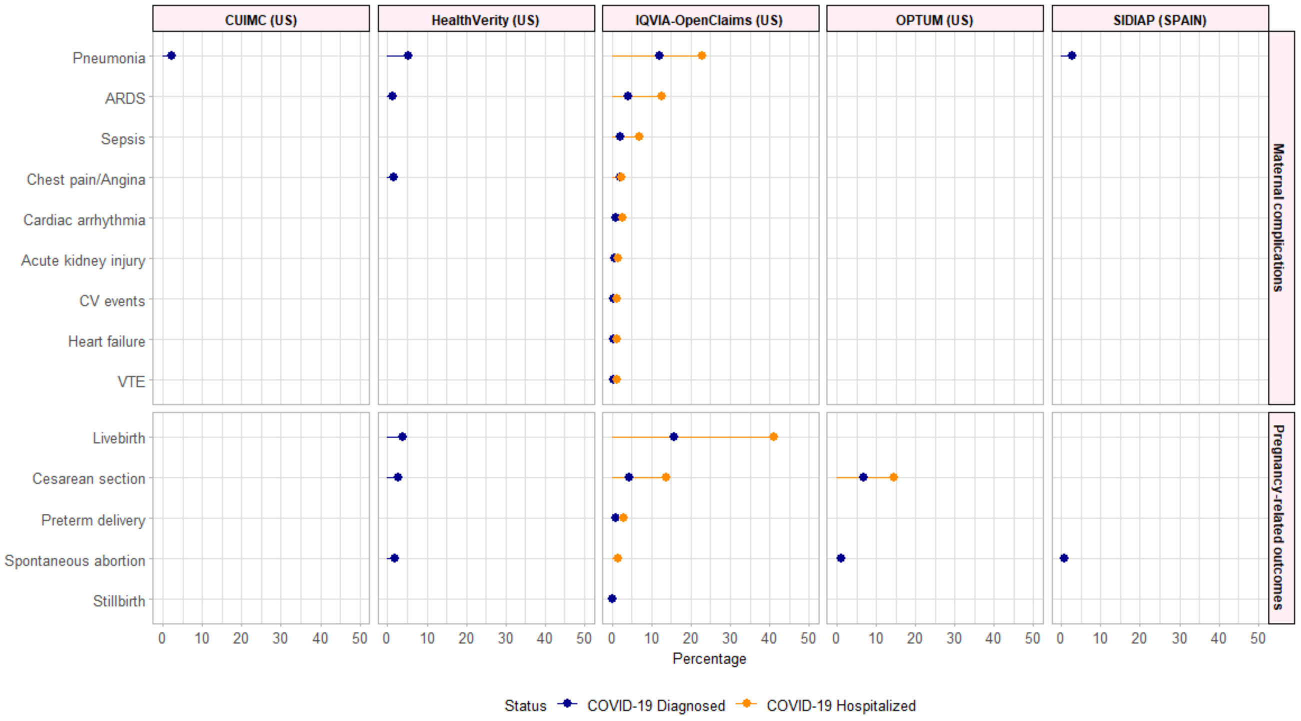


**Supplementary Figure 2b. Maternal complications and pregnancy-related outcomes amongst women diagnosed with COVID19 versus diagnosed with influenza across all databases**

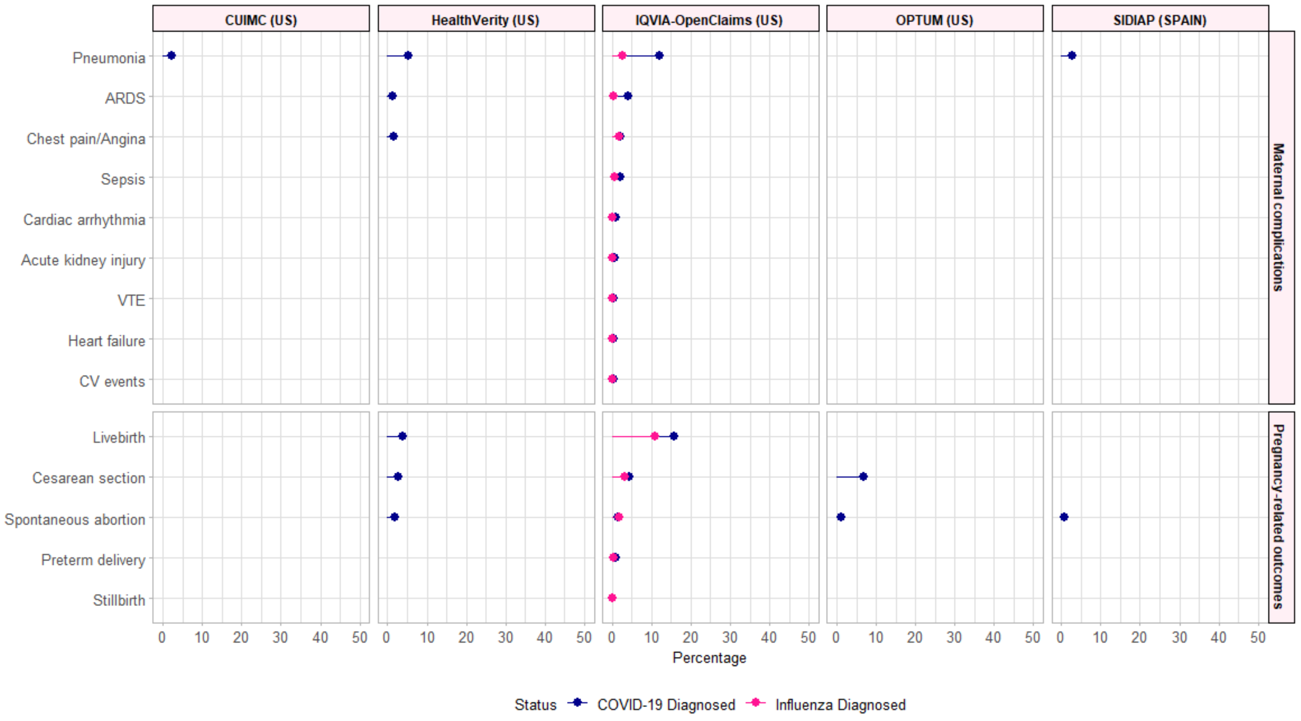
